# Risk Prediction for ALS using Semi-competing Risks Models with Applications to the ALS Natural History Consortium Dataset

**DOI:** 10.1101/2024.11.26.24317991

**Authors:** Andres Arguedas, David Schneck, Erjia Cui, Annette Xenopoulos-Oddsson, Ximena Arcila-Londono, Christian Lunetta, James Wymer, Nicholas Olney, Kelly Gwathmey, Senda Ajroud-Driss, Ghazala Hayat, Terry Heiman-Patterson, Federica Cerri, Christina Fournier, Jonathan Glass, Alex Sherman, David Walk, Mark Fiecas

## Abstract

**Background and objectives:** Important landmarks in progression of ALS can occur prior to death. Predictive models for the time these occur can assist in clinical trial design and personal patient planning. We propose a predictive model, using a semi-competing risks modeling approach, for five important disease progression landmarks in ALS.

**Methods:** Longitudinal data on 1,508 participants from the ALS Natural History Consortium were used, including the ALSFRS-R score collected at different clinic visits and baseline patient characteristics. A semi-competing risks modeling approach was used to study the time to certain disease progression landmarks accounting for the possibility of death. Specifically, time to gastrostomy, use of NIV, continuous use of NIV, loss of speech, and loss of ambulation were chosen and modeled individually. To measure the predictive capabilities of the model, the integrated Brier score was computed for each model using cross-validation for the NHC data. Data from Emory University were used for external validation of the models.

**Results:** We present model results using gastrostomy as the intermediate outcome. Similar trends in disease progression groups were found across all model pathways. The sub-scores for speech and swallow, and respiratory function, as well as diagnostic delay, were the most important covariates. Predictive metrics in both internal and external validation are presented across all models and for different pathways.

**Conclusions:** Semi-competing risks modeling is a flexible approach to studying disease progression. Our data show that this modeling approach can be used to inform clinical trial modeling and people living with ALS.

## Introduction

Amyotrophic lateral sclerosis (ALS) causes muscle weakness, fatigue, dysarthria, dysphagia, respiratory failure, loss of mobility, and death. The median survival time is 2-4 years, with less than 10% of people living with ALS surviving past 10 years(1). In addition to time to death, disease progression can be measured as time to common events (“landmarks”) in functional progression. These landmarks include gastrostomy, use of non-invasive ventilation, loss of speech, and loss of ambulation. Because ALS presents in different regions of the body in different individuals, whereas death from ALS is almost always due to ventilatory failure, time to these events, or whether they occur at all prior to death, varies greatly among people living with ALS.

There is currently no treatment that results in stabilization or improvement for most people living with ALS. One of the barriers to therapeutic development in ALS is the wide variability and limited predictive information in disease progression. Predictive modeling of times to events would aid in developing models for clinical trial development, evaluation of real-world evidence, and personal planning. Specifically, any approach to model and predict intermediate events should account for death as an additional terminal event, as well as being able to include different predictions for after a person experiences intermediate events. Semi-competing risks models (or illness-death models) use a modeling approach that explicitly accounts for the fact that a person can die after experiencing an intermediate event, but not vice versa. Therefore, these models can be used to study what characteristics affect different parts of a person’s disease progression. For example, a certain characteristic can be more strongly associated with the intermediate event than death, or it might only be associated with death. Hence, this modeling approach allows us to study the time to intermediate events or death, but also how they relate and how outcomes change after experiencing the intermediate event. This information can be used both to better understand the intermediate events and disease progression as a whole, and for patients and clinicians to plan accordingly for the possibility of the occurrence of intermediate events. These types of models have been previously used in other areas, including obstetrical(2), aging(3), and cancer studies(4,5).

Our objective is to adapt the semi-competing risks modeling framework for modeling ALS progression. At a population level, the ability to describe and predict the probability of these events occurring at a certain time point can be used to better inform the design of clinical trials in ALS. At an individual level, this would enhance personal planning for people living with ALS, their caregivers, and clinicians.

## Methods

### Data

The ALS Natural History Consortium (NHC) dataset was utilized. The ALS NHC dataset contains information on people living with ALS abstracted from multidisciplinary ALS clinic care records. As of March 2nd 2023, the ALS Natural History Consortium dataset contained information from a total of 1,977 participants seen at 10 participating centers. The dataset represents a combination of baseline characteristics and longitudinal disease progression data that are obtained in the course of routine clinical practice, including the ALS Functional Rating Scale - Revised (ALSFRS-R)(6). The ALSFRS-R score is an instrument used to measure disease progression in people with ALS across time. It consists of 12 questions with scores from 0 to 4, ranging from a severe limitation to perform a certain activity to no limitation at all, respectively. The total ALSFRS-R score can be obtained by summing the score from each question, and thus ranges from 0 to 48. The ALSFRS-R can further be separated into 4 domains: 1) Speech and Swallow (questions 1-3), 2) Fine Motor (questions 4-6), 3) Gross Motor (questions 7-9), and 4) Respiratory Function (questions 10-12). Since each of the 4 domains consists of 3 questions, the respective subscore can be obtained by summing the corresponding questions for each subscore, for a range from 0 to 12. In addition to the ALSFRS-R, additional baseline characteristics included for this analysis are age, sex, site of onset (limb or bulbar), and use of riluzole. Finally, the time between symptom onset and diagnosis was derived as the diagnostic delay for each person.

The intermediate events of interest used in this analysis were gastrostomy, use of non-invasive ventilation (NIV), continuous use of NIV, loss of speech, and loss of ambulation. The dataset provides the date of gastrostomy. Use of NIV, continuous use of NIV, loss of speech, and loss of ambulation were abstracted from the ALSFRS-R data, which were obtained a median of every 98 days. To estimate the actual date, we use the first observed date in which the score for a corresponding question in the ALSFRS-R was below a certain threshold.

The data used to determine that a person experienced each intermediate event and corresponding threshold were: 1) for use of NIV, indicating intermittent use of BiPAP or worse in question 12 (*“Respiratory insufficiency”*), 2) for continuous use of NIV, indicating continuous use of BiPAP during the night and day or worse in the same question, 3) for loss of speech, indicating loss of useful speech in question 1 (*“Speech”*) and 4) for loss of ambulation, indicating non ambulatory functional movement or worse in question 8 (*“Walking”*). The last date at which information from participants was obtained was determined as the date of their last ALSFRS-R visit. This was used for potential censoring times. Participants who had only one observed ALSFRS-R score after diagnosis were also excluded. Some participants had a gastrostomy before diagnosis of ALS and thus were subsequently excluded only from the analytic sample used for the models with the gastrostomy outcome.

To perform an external validation of our model, an ALS clinic dataset separate from ours was obtained from Emory University (“Emory dataset”). This dataset included the same variables listed above and went through the same exclusion criteria/restrictions as those applied to the NHC data. Edaravone was not available in this dataset and was thus not included as a covariate in our models. A summary of the baseline characteristics and disease progression variables for this dataset are presented in the Supplements as Table S.1.

All participants provided written informed consent to be included in the database. Current IRB numbers: University of Minnesota (UMN) sIRB: 00019204; Istituti Clinici Scientifici Maugeri: 2687CE; and Centro Clinico NeMO: 247-052017. Prior IRB numbers: UMN: 1501M61381; Northwestern: STU00209860; Saint Louis University: 28018; Temple: 25661; Virginia Commonwealth University: HM20009645; University of Florida: IRB201701910; Henry Ford: 10105; and Providence: 16-123A. Data from Emory University collected through the Clinical Research in ALS Study (CRiALS), IRB: 00078771.

### Data Availability Statement

The dataset used for this study can be made available upon reasonable request for research purposes.

### Modeling approach

A semi-competing risks model(7) was used to account for the possibility of an intermediate event occurring before death. Thus, there are two possibilities for every participant: 1) they experienced the intermediate event, or 2) they did not experience the intermediate event. For those who experienced the intermediate event, we can further detail what happened to them before and after the intermediate event. For those who did not experience the intermediate event, death may occur before the event happens, or they may be censored. Hence, three possible pathways for disease progression can be modeled simultaneously using a semi-competing risks model: 1) death before experiencing the intermediate event, 2) experiencing the intermediate event without death during the period of observation, and 3) death after the intermediate event during the period of observation. These pathways, as well as examples of disease progression across time, are presented in Figure 1.

**Figure 1.**
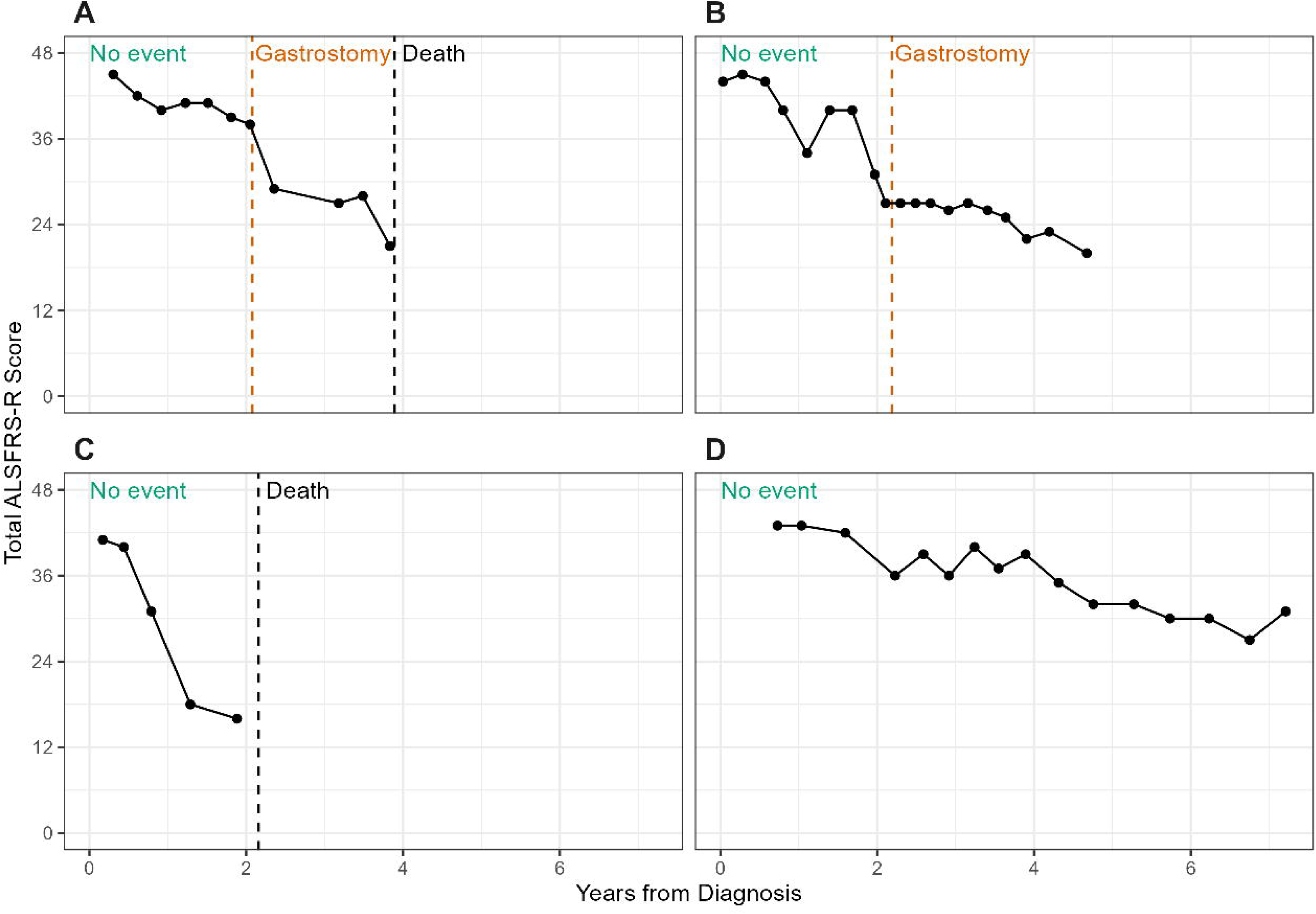
Examples of possible disease progression pathways, based on ALSFRS-R scores across time and time to important events. (A) A person who died after experiencing an intermediate event (pathway 3). (B) A person who has experienced an intermediate event, but not yet death (pathway 2). (C) A person who died before experiencing an intermediate event (pathway 1). (D) A person who has not yet experienced either an intermediate event or death.

Five models were created, one for each of the intermediate events (gastrostomy, use of NIV, continuous use of NIV, loss of speech, and loss of ambulation), with each model studying the three possible disease pathways simultaneously using hazard functions. All the variables listed in the Data section were included in each of these models. Since multiple measurements of the ALSFRS-R score could be taken longitudinally over time, a summary measure of the change in the subscores was used. Specifically, a mixed effects model with normal errors was computed using each of the ALSFRS-R subscores as the response variables, with a fixed effect for time, and a random intercept and slope for each participant. The predicted random intercept and slope for each participant were used as covariates in the semi-competing risks models. Additionally, the random intercepts were truncated to take a value between 0 and 12, corresponding to the possible values of the ALSFRS-R subscores.

All covariate effects were independent across pathways, and a Semi-Markov assumption was chosen for all further modeling purposes. This assumes that the hazard of death after the intermediate event depends on the time spent in this pathway, as well as the time when the intermediate event was observed. A Weibull proportional hazards model was used to determine both the baseline hazards as well as the functional relations between the hazards and covariates.

Once the models were fit, the predicted event times of each participant were obtained for each pathway and model, and their corresponding quintiles were calculated. Participants were categorized into five groups of disease progression using the quintiles from the path from “No Event” to “Death”: 1) very slow, 2) slow, 3) intermediate, 4) fast, and 5) very fast. These categories were created to aid in visually representing the results of the model, as well as relationships between the survival probabilities of different pathways.

### Predictive Metrics

The predictive capabilities of all models was compared using the Integrated Brier Score (IBS). In this case, the Brier Score (BS) for each model was calculated at 0, 6, 12, 24, 60, and 120 months, and subsequently used to obtain the IBS. The IBS can take values between 0 and 1, with 0 representing perfect predictive accuracy (at every time point, the model perfectly predicts the survival probability for each participant), and 1 representing completely incorrect predictions (at every time point, the model incorrectly predicts the survival probability for each participant). A value of 0.25 or lower for the IBS can be used as a rule of thumb to indicate good predictive capabilities. Since a semi-competing risks model was used for all five intermediate events, the IBS calculation was done for the three different pathways separately in each model. For the NHC data, 10-fold cross-validation was used to assess the predictive capabilities of the models. Hence, the IBS for each fold was averaged to obtain a final value for the IBS of each model for each different path. Based on the model fit with all of the NHC data, the IBS was calculated when performing predictions using the Emory dataset.

### Software

All analyses were done using R version 4.2.3, using the *tidyverse*(9) and *mstate*(10) packages for data cleaning, wrangling, and plotting. For modeling, the *survival*(11), *lme4*(12), and *SemiCompRisks*(13) packages were used. The *riskRegression*(14) package was used to compute the IBS. We also developed a Shiny companion application to interactively present the results for different models, which also allows for users to provide inputs of individual characteristics and subsequently obtain predictions of survival probabilities based on the models presented in this manuscript.

## Results

The ALS NHC dataset provided for this work included data from 1,977 participants, of whom 1,508 were used for our modeling. See Figure 2 for the CONSORT diagram. Cohort characteristics are available in Table 1. The Emory dataset included 1,288 participants, and cohort characteristics are presented in Table S.1.

**Figure 2.**
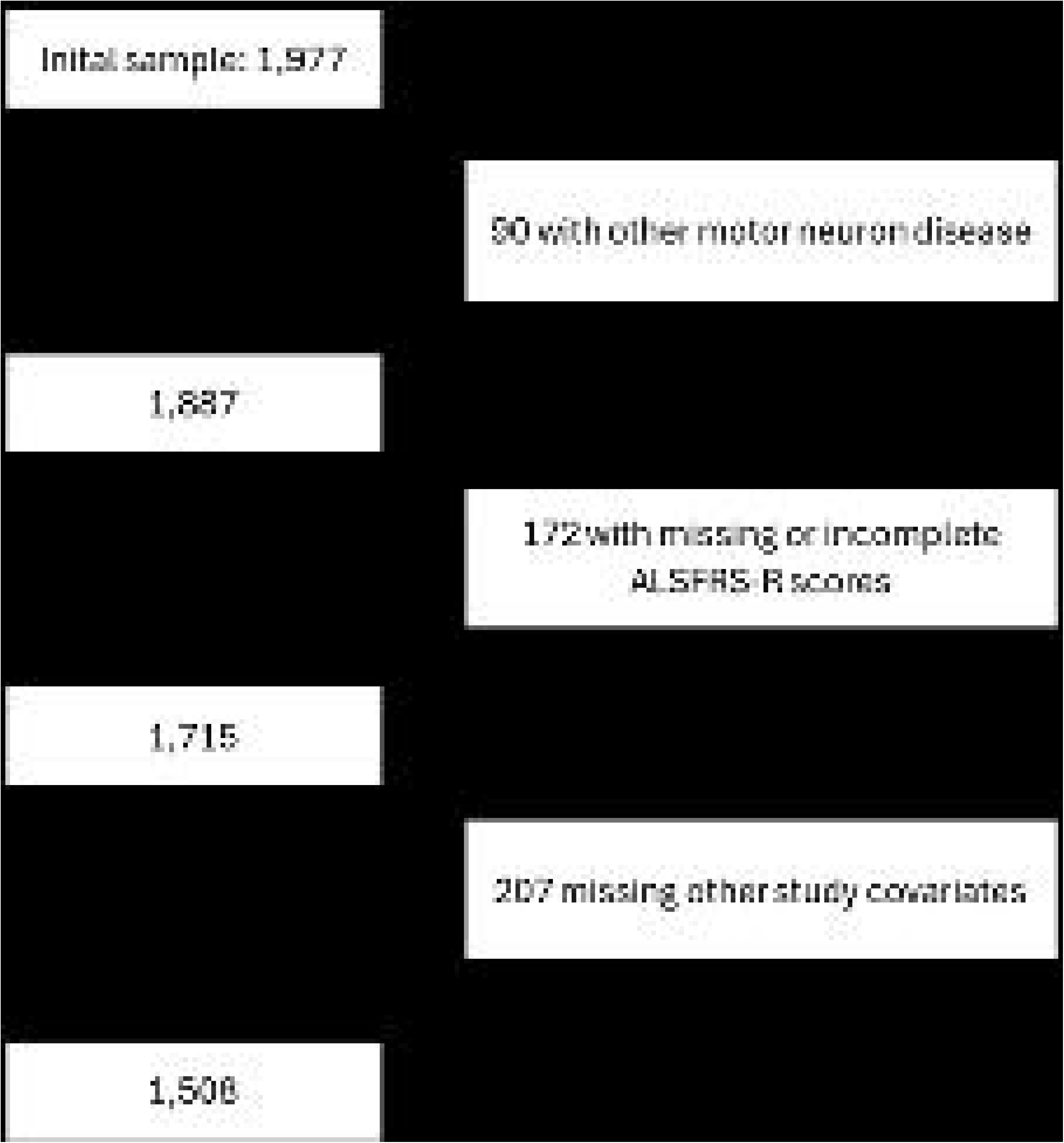
Eligibility criteria diagram for the analytic sample

**Table 1:** Cohort characteristics for participants in the analytic sample from the NHC data.

| Characteristic | N = 1,508<sup>1</sup> |
| --- | --- |
| Male | 889 (59.0%) |
| Limb onset site | 1,020 (67.6%) |
| Age at Onset (years) | 62.38 (54.7, 69.2) |
| Age at Diagnosis (years) | 64.00 (56, 71) |
| Uses Riluzole | 1,184 (78.5%) |
| Number of ALSFRS visits | 4 (3, 7) |

<sup>1</sup> n (%); Median (IQR)

Due to the considerable number of intermediate events analyzed, in this section we primarily present the results for the model using gastrostomy as the intermediate event. Because 34 participants had gastrostomies before their diagnosis of ALS, the sample size for the model containing this intermediate event was 1,474.

The median time to death without experiencing a gastrostomy is 4.25 years (95% CI: 3.66-4.93), for experiencing gastrostomy it is 5.44 years (95% CI: 4.95-7.38), and for death after experiencing a gastrostomy it is 1.47 years (95% CI: 1.28-1.70). From these summaries, we see that the behavior of each of the three disease progression pathways is different, and thus studying them separately using a semi-competing risks model, as well as allowing for different baseline hazards and covariate effects, may be useful, since these results cannot be obtained using a single simple Cox proportional hazards model.

Figure 3 presents the estimated survival curves for each disease progression pathway obtained from our semi-competing risks model and for each of the five different disease progression groups defined using the predicted median survival time from our model.

**Figure 3.**
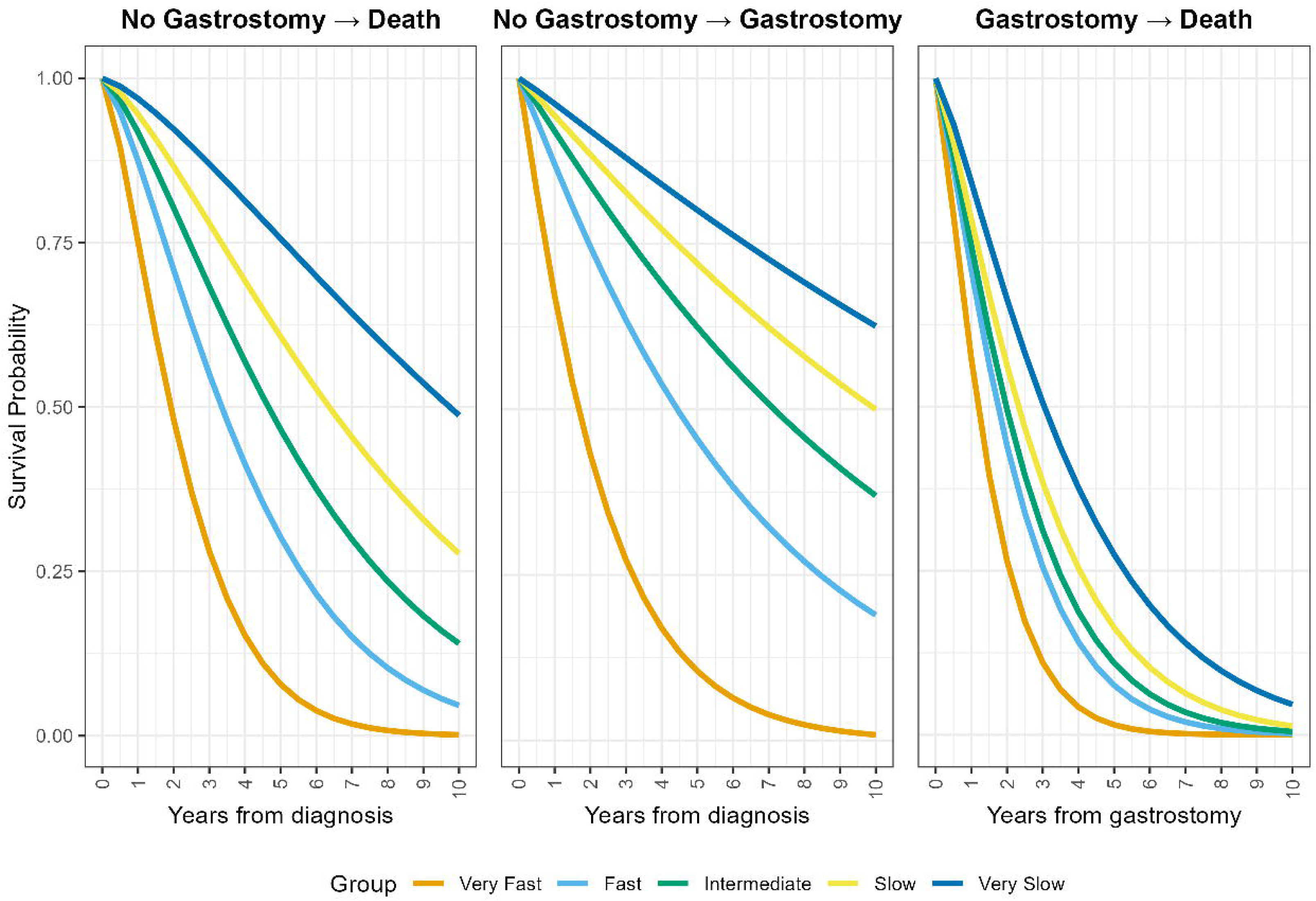
Estimated survival curves for each of the three different disease progression pathways, when using the median predicted event times for each of the progression groups. Progression groups were created based on the quintiles of the predicted event times for the time to death without experiencing a gastrostomy pathway.

Figure 4 presents a summary of the predicted survival curves for a sample of three participants from each of the five progression groups, while Table 2 presents the values of the covariates and characteristics of the participants presented in Figure 4.

**Figure 4.**
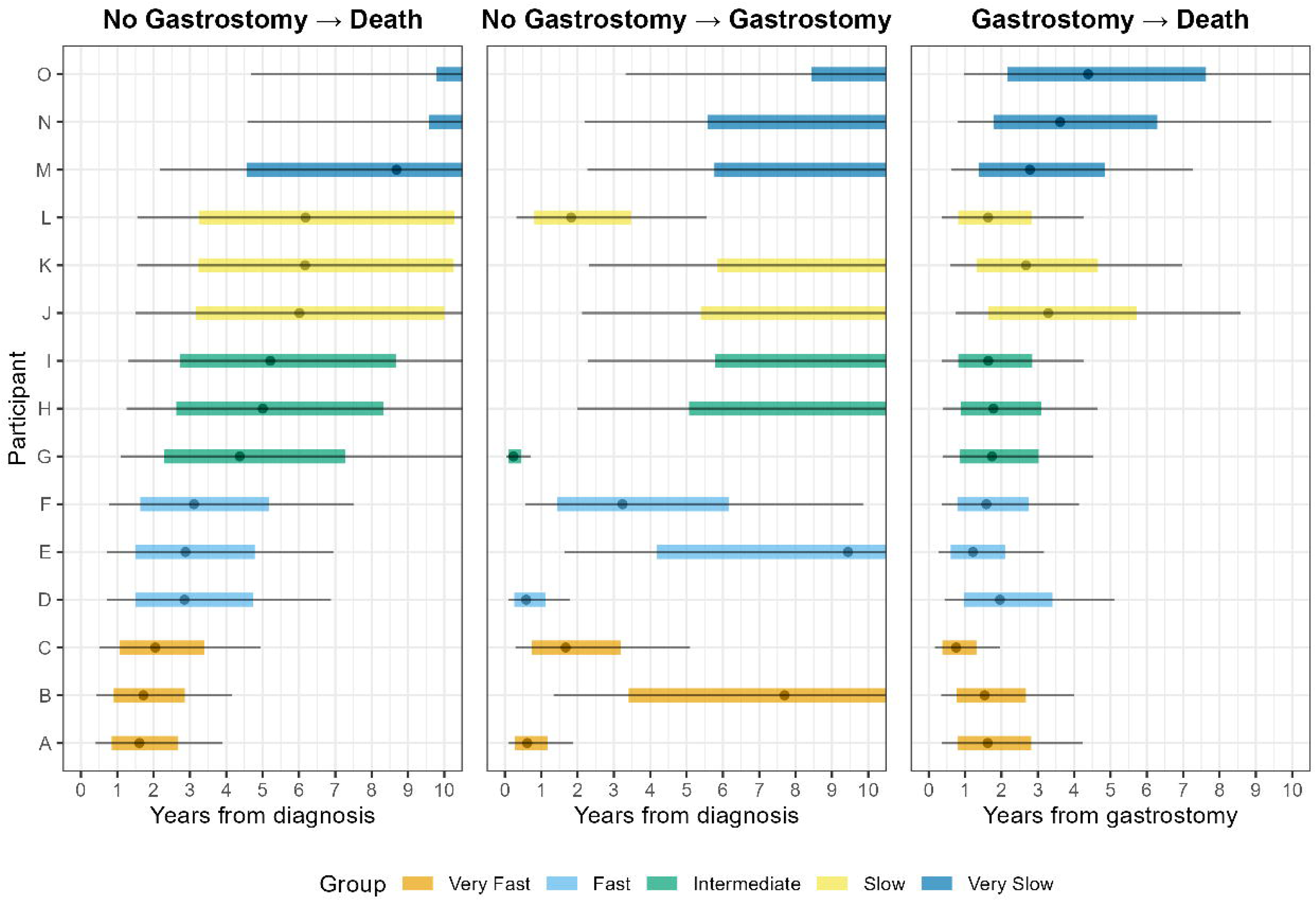
Summary of the predicted survival probability curves for a sample of participants in each disease progression group, according to the pathway. The left whisker represents the time point at which the survival probability crosses 0.90, while the left border of the box corresponds to 0.75, the dot in the middle to 0.50, the right border of the box to 0.25, and the right whisker to 0.1

**Table 2:** Characteristics and derived covariates of a sample of participants in each disease progression group.

| Participant | Progression Group<sup>1</sup> | Age<sup>2</sup> | Sex<sup>2</sup> | Site of Onset | Uses Riluzole | Diagnostic Delay (years) | Random Effects - ALSFRS-R subscores |  |  |  |  |  |  |  |
| --- | --- | --- | --- | --- | --- | --- | --- | --- | --- | --- | --- | --- | --- | --- |
|  |  |  |  |  |  |  | Speech & Swallow |  | Fine Motor |  | Gross Motor |  | Resp. Function |  |
|  |  |  |  |  |  |  | Baseline Score | Rate of Change | Baseline Score | Rate of Change | Baseline Score | Rate of Change | Baseline Score | Rate of Change |
| O | VS | 50-59 | - | Limb | No | 6.73 | 9.44 | -0.02 | 7.08 | 0.00 | 5.79 | -0.04 | 10.21 | 0.09 |
| N | VS | 60-69 | - | Limb | No | 5.75 | 8.60 | -0.01 | 10.35 | -0.02 | 6.95 | -0.01 | 10.39 | 0.01 |
| M | VS | 40-49 | - | Limb | Yes | 0.85 | 10.86 | 0.00 | 10.79 | -0.14 | 11.77 | -0.23 | 11.61 | -0.05 |
| L | S | 70-79 | - | Bulbar | Yes | 0.59 | 5.56 | -0.10 | 12.00 | -0.14 | 12.00 | -0.14 | 11.49 | 0.01 |
| K | S | 50-59 | - | Limb | Yes | 1.34 | 12.00 | -0.05 | 7.84 | -0.13 | 7.66 | -0.11 | 12.00 | -0.17 |
| J | S | 50-59 | - | Limb | Yes | 0.70 | 12.00 | -0.08 | 5.61 | 0.02 | 5.37 | -0.09 | 12.00 | -0.19 |
| I | I | 80-89 | - | Limb | No | 7.17 | 12.00 | -0.14 | 5.29 | -0.09 | 4.05 | -0.08 | 7.86 | -0.15 |
| H | I | 70-79 | - | Limb | Yes | 1.83 | 10.85 | 0.02 | 9.98 | -0.03 | 9.85 | -0.29 | 10.46 | -0.10 |
| G | I | 50-59 | - | Bulbar | No | 0.91 | 3.84 | -0.54 | 4.95 | 0.04 | 5.33 | -0.16 | 8.70 | -0.15 |
| F | F | 70-79 | - | Limb | Yes | 3.00 | 8.94 | -0.19 | 7.82 | -0.22 | 6.76 | -0.19 | 9.70 | -0.21 |
| E | F | 70-79 | - | Limb | No | 1.05 | 9.64 | 0.00 | 9.16 | -0.34 | 4.20 | -0.07 | 11.84 | -0.10 |
| D | F | 60-69 | - | Bulbar | Yes | 0.33 | 6.94 | -0.35 | 6.53 | -0.18 | 5.85 | -0.04 | 8.00 | -0.29 |
| C | VF | 60-69 | - | Limb | Yes | 0.15 | 12.00 | -0.53 | 8.28 | -0.47 | 6.42 | -0.31 | 12.00 | -0.24 |
| B | VF | 60-69 | - | Limb | Yes | 0.54 | 11.85 | 0.01 | 9.64 | -0.37 | 6.40 | -0.31 | 10.53 | -0.41 |
| A | VF | 60-69 | - | Bulbar | No | 0.63 | 5.79 | -0.29 | 2.80 | -0.20 | 1.99 | -0.14 | 6.10 | -0.21 |

<sup>1</sup> Disease progression groups are: VS - very slow, S - slow, I - intermediate, F - fast, VF - very fast
<sup>2</sup> To preserve the anonymity of participants, age was put into deciles and sex was removed from presentation. Sex and precise age were used to fit the model

### Predictive Metrics

The predictive capabilities of each model were measured using the IBS. Table 3 presents the IBS obtained using cross-validation for the NHC data, as well as for the predictions from the Emory data, for each intermediate event model, by the different disease progression pathways.

**Table 3:** IBS for each of the five outcomes of interest, by the three different disease progression pathways.

| Intermediate Event | Data | IBS for Pathway |  |  |
| --- | --- | --- | --- | --- |
|  |  | No Intermediate Event to Death | No Intermediate Event to Intermediate Event | Intermediate Event to Death |
| Gastrostomy | NHC | 0.159 | 0.123 | 0.190 |
|  | Emory | 0.207 | 0.132 | 0.180 |
| NIV | NHC | 0.266 | 0.132 | 0.181 |
|  | Emory | 0.281 | 0.162 | 0.159 |
| Continuous use of NIV | NHC | 0.150 | 0.079 | 0.222 |
|  | Emory | 0.185 | 0.070 | 0.185 |
| Loss of Ambulation | NHC | 0.179 | 0.124 | 0.208 |
|  | Emory | 0.205 | 0.139 | 0.190 |
| Loss of Speech | NHC | 0.161 | 0.064 | 0.183 |
|  | Emory | 0.199 | 0.070 | 0.172 |

The IBS between the two datasets seem to be consistent, although they vary between intermediate events and pathways. In general the IBS is slightly lower (more predictive) for the NHC data when predicting either death without experiencing an intermediate event or when predicting experiencing an intermediate event compared to experiencing death after an intermediate event. On the other hand, when predicting death after an intermediate event, the Emory data has a lower IBS for all intermediate events. For most events except NIV, the IBS for predicting death after an intermediate event seems to be the highest, followed by predicting death without experiencing an intermediate event, and is lowest (most predictive) for predicting an intermediate event without death.

## Discussion

The semi-competing risks approach allows us to predict intermediate events separately from death and to predict death after an intermediate event has occurred. Thus, in contrast with using a composite endpoint(15,16), probabilities of the intermediate event occurring over time can be given for participants, separately from death. Compared to other predictive models for ALS(17), our modeling approach takes into account five different intermediate events, although more could be included, and thus important information on disease progression can be modeled more thoroughly. For example, time to advancing in ALS staging, such as Kings’(18) or MiTOs(19), could be modeled. This can empower and inform people living with ALS as well as their caretakers and clinicians in making important personal and clinical decisions.

A semi-competing risk modeling approach efficiently models overall disease progression while allowing for differences between possible pathways. The model allows for different associations between pathways, giving greater importance to certain variables on different outcomes. This is further demonstrated by the model’s predictive capabilities. Although the model has good overall predictive power, some pathways have better performance than others, and this can also vary across different outcomes. This indicates that it is not sufficient to model all pathways together, or to assume that different outcomes have similar results, further supporting the use of a semi-competing risks model.

Our results were consistent between the internal and external validation datasets. This motivates a broader application for this model in new and different cohorts, both for validation, but also for use in prediction. For readers interested in incorporating dynamic predictions within a landmarking framework for analyzing time-to-event outcomes in ALS, the work of Schneck et. al. provides valuable insights.

Our modeling has limitations. First, while the ALS NHC records include disease data from the time of diagnosis or before, 62.7% of participants were enrolled as prevalent cases, defined by the consortium as an enrollment date more than 90 days after the date of diagnosis. Hence participants with short disease duration may be underrepresented. Edaravone use was not included in our model as that information was not available in our validation dataset. Additionally, although time to gastrostomy was recorded from medical visits, all other intermediate events are based on ALSFRS-R scores. Therefore, the exact date is not known for these intermediate events.

### Next steps

As more variables and biomarkers are obtained our model will be refined to include those which demonstrate predictive utility. We will further extend this to a dynamic framework that allows for time-varying effects in this semi-competing risks modeling framework.

## Acknowledgements

This project was supported by the FDA’s Office of Orphan Products Development under grant number FD-R01-0007630. Its contents are solely the responsibility of the authors and do not necessarily represent the official views of the FDA nor FDA’s Office of Orphan Products Development.

## Disclosure Statement

XAL reports the Trial Capacity Award Recipient 2022, ALS Association Grant; CL received compensation for scientific consulting from Mitsubishi Tanabe Pharma Europe and Biogen, his research has been funded by grants from ALSA, ARISLA, the Italian Ministry of Health, and the Italian Ministry of Research, his work has been partially supported by the Italian Ministry of Health’s RC program; JW reports MT Pharma for grant support; KG has received consulting honoraria from Alexion, Argenx, Amgen and UCB, remotely received speaking honoraria from Argenx and Alexion; SAD reports research support from Biogen, Amylyx pharmaceuticals, Mitsubishi Tanabe Pharma America, Alnylam, Novartis, Sanofi, reports personal consulting fees from Biogen and Amylyx pharmaceuticals, served as a paid educational presenter for Biogen; GH reports support from Speaking Bureau, Alexion, Argenx, MTPA; THP reports support from Medical Advisory Board for Mitsubishi Tanabe Pharma America, Amylyx, Novartis, Clinical trial funding: Amylyx, Novartis, Mitsubishi Tanabe Pharma, Healey Platform Trial, Clinical Research Funding: Amylyx, Mitsubishi Tanabe Pharma America; CNF reports consulting fees from Novartis, Roon, and QurALIS; JDG reports funding from NINDS and NIA, Consulting for Biogen, NuraBio, Aruna Bio, Third Rock; AS has received grants and contracts for clinical research projects sponsored by FDA, NIH/NIA, NIH/NINDS, The ALS Association, and ALS Finding a Cure Foundation as well as study support from MTPA, Biogen, and Amylyx; DW reports grant support from FDA’s Office of Orphan Products Development and consulting fees from Biogen, Mitsubishi Tanabe Pharma America, and Amylyx. All other authors have no relevant disclosures to report.

## Supplements

### Appendix 1 - Distribution of data from Emory University

The standardized mean difference (SMD)(20) is a metric used to quantify the differences in variables between two groups. Originally defined for use only with numeric variables, further developments extended the SMD to be used with binary variables(21). Various authors suggest rules of thumb for categorizing differences based on the SMD. Generally, an SMD of 0 indicates that the two groups have identical distributions for that variable, while a value smaller than 0.1 is usually associated with negligible differences, between 0.1 and 0.2 as small differences, and greater than 0.2 as noticeable differences. Table S.1 presents the distribution of the variables of interest for Emory University’s dataset, as well as for the NHC data, using the SMD to quantify the differences between them for each variable.

Riluzole use shows a particularly large difference between both cohorts, with a larger proportion of participants in the NHC using riluzole. There are also differences in the site of onset, age at onset and at diagnosis, as well as the number of ALSFRS visits, although these are smaller. Finally, both cohorts have a very similar proportion of males and females.

---

**Table S.1:** Cohort characteristics for participants in the analytic sample from Emory University’s ALS data compared to those from the Natural History Consortium DatasetFigure Captions.

| Characteristic | Emory (N = 1,288)<sup>1</sup> | NHC (N = 1,508)<sup>1</sup> | SMD<sup>2</sup> | p-value<sup>3</sup> |
| --- | --- | --- | --- | --- |
| Males | 760 (59.0%) | 889 (59.0%) | 0.001 | >0.99 |
| Limb onset site | 947 (73.5%) | 1,020 (67.6%) | 0.13 | <0.001 |
| Age at Onset (years) | 60.0 (51.0, 68.2) | 62.4 (54.7, 69.2) | 0.22 | <0.001 |
| Age at Diagnosis (years) | 61.5 (53.0, 69.5) | 64.0 (56.0, 71.0) | 0.21 | <0.001 |
| Uses Riluzole | 443 (34.4%) | 1,180 (78.5%) | 0.99 | <0.001 |
| Number of ALSFRS visits | 3 (1, 5) | 4 (3, 7) | 0.24 | <0.001 |

<sup>1</sup> n (%); Median (IQR)
<sup>2</sup> Standardized Mean Difference
<sup>3</sup> Pearson's Chi-squared test

